# Predictive Classification of IBS-subtype: Performance of a 250-gene RNA expression panel vs. Complete Blood Count (CBC) profiles under a Random Forest model

**DOI:** 10.1101/2021.08.31.21262766

**Authors:** Jeffrey Robinson

**Author notes:** LinkedIn: https://www.linkedin.com/in/robinsonjmevol/. ResearchGate: https://www.researchgate.net/profile/Jeffrey_Robinson4.

## Abstract

In this experiment, an R-script was developed to select the best performing machine learning (ML) predictive classification algorithm for IBS-subtype, and compare the performance of two datasets from the same clinical cohort – 1) The Complete Blood Count (CBC) results, and 2) A 250-gene Nanostring expression panel run on RNA from the “Buffy Coat” fraction. This publicly available data was compiled from open-source repositories and previously published supplementary data. Column labels were reformatted according to “tidy-data” standards. NA values in the data were imputed based on the mean value of the data column. Subject groups included Control (ie. healthy), IBS-D (diarrhea predominant), and IBS-C (constipation predominant) subtypes. These groups had unequal numbers in the original study, and so random re-sampling was used to make the group numbers equal for downstream linear regression-based analyses. The data was randomly split into training and validation subsets, and 5 classification algorithms were tested. Random Forest was clearly the best performing algorithm for both CBC and gene expression panel data, generally with >95% predictive accuracy, without additional tuning. The 250-gene RNA expression panel performed somewhat better than the CBC profile under a Random Forest model, however the CBC profiles had only 13 predictor variables vs. the 250 of the RNA expression panel. Some artifacts may result from the duplication of IBS-D and IBS-C rows from to the group-size balancing method, and so larger and more comprehensive datasets will be obtained for a follow-up analysis. The R-script and reformatted data are published as supplementary material here, and as a component of the ‘AnalyzeBloodworkv1.2’ GitHub repository.

## Introduction

Differential diagnosis of GI diseases with similar symptom profiles has been an active area of clinical research. IBS, gastroenteritis, IBD, and other conditions present with similar symptoms including diarrhea, constipation, and gastro-abdominal pain, and biomarker profiling strategies incorporating symptom-based inventories have proven accurate in differential diagnosis of IBS vs other pathologies with similar symptom profiles. (Manning et al. 1978)

Molecular and cellular biomarker strategies ideally provide fast, accurate, and cheap diagnosis. An example of a cell-count/cell-morphology based test panel is the standard (and venerable) Complete Blood Count with differential (CBC-D). This flow-cytometry test counts Neutrophils, Macrophages, Lymphocytes, Granulocytes, Red Blood Cells with additional parameters (ie. ESR, HCT, MCH), Platelets count and volume (MPV). These datapoints are sufficient to narrow down a variety of differential diagnoses. (Van Leeuwen et al. 2006)

We previously showed that some of the CBC-D exhibited some statistical association with IBS-subtype in a clinical natural history cohort (Robinson et al. 2019). However, this analysis remained flawed by uneven group sizes and significant missing data from the original study, which violates assumptions for linear regression and other statistical methodology. We therefore sought to improve the analysis by implementing common statistical methods such as imputation of missing data points and random re-sampling to balance the group sizes.

White Blood Cell (“Buffy Coat”) fractions were also collected, and RNA expression profiles were developed for the same clinical cohort using a 250-gene Nanostring gene expression panel (Robinson 2019). Both the raw and normalized Nanostring expression data and gene list and probe details of the 250-gene Nanostring probe panel “ImmunoGC” are available in the NCBI Gene Expression Omnibus (GEO) database. (Expression data: https://www.ncbi.nlm.nih.gov/geo/query/acc.cgi?acc=GSE124549; Probe panel: https://www.ncbi.nlm.nih.gov/geo/query/acc.cgi?acc=GPL25996). Briefly, the probe panel contains genes associated with cell-type specific immune cell genes, canonical adaptive/innate immune pathway genes, miRNA pathway genes, and others.

In the following report, we take these datasets and compare their performance of several different machine learning classification algorithms, using the R-package “Caret” :

1. Linear Discriminant Analysis (LDA),
2. Classification and regression tree (CART),
3. K-nearest neighbors (KNN),
4. Support Vector Machine (SVM), 5) Random Forest (RF).

Based on the results of this model selection, we then took the best performing algorithm (Random Forest), and compared the accuracy of CBC-D-based models vs. gene-expression-based models using confusion matrix.

Complete results are presented below. R-language code for replicating this analysis is provided as supplementary methods. “tidy”-formatted CBC-D and Nanostring data files are provided as supplementary data. This code and data content are also found within the updated “AnalyzeBloodwork1.2” GitHub repository.

## Methods

### A. R-code

R-code was developed to perform these analyses in an automated manner. In this version 1 of the pre-print, the supplementary .zip file contains the script “IBSclassification.R” and .csv-format data (“RobinsonEtAl_Sup1.csv” and “mRNAnorm.csv”, for CBC-D and gene expression data, respectively) within the proper file structure. Once the file is unzipped, the script “IBSclassification.R” can be opened in R-Studio and run line-by-line without modification to obtain identical results. The script was developed in RStudio v1.3.1073, using R version 4.0.2 (2020-06-22) on Windows 10 (x86_64-w64-mingw32). Running the code with other versions of R or R-Studio cannot be guaranteed to run error-free.

The script will perform the following for the CBC-D and RNA expression data, respectively:

1. Install and load required packages,
2. Load and pre-process the data files,
3. Impute missing data and balance group sizes through random re-sampling,
4. Split data into “training” and “validation” subsets,
5. Train the classification models using the 5 algorithms mentioned above,
6. Display a dotplot showing the predictive performance of each algorithm,
7. Estimate the “skill” of the Random Forest models with a confusion-matrix analysis,
8. Generate error-rate by number of trees generated plots.

The code and modified data for this project is provided as **<u>Supplementary Material 1</u>**. The code and results are also incorporated into the GitHub repository AnalyzeBloodwork1.5 software (Robinson, 2021; https://github.com/PhyloGrok/AnalyzeBloodwork).

### B. Machine-Learning model selection workflow

The machine-learning in R workflow presented by Brownlee (2016) was adapted for this project, with a more formalized presentation of the workflow found in Brownlee (2020). R “caret” package provided the primary machine learning functions used for this code (Kuhn 2021).

### C. Random Forest model and visualizations

Briefly, the random forest algorithm generates a large number of random decision-trees, and then refines and selects the most accurate combined set of decision trees as an “ensemble” for classification decisions (Yiu, 2019).

Additional help for coding the random forest diagnostic plots came from the following web resources: https://stackoverflow.com/questions/39636186/plot-decision-tree-in-r-caret https://stats.stackexchange.com/questions/51629/multiple-curves-when-plotting-a-random-forest

## Results

### Algorithm selection (**Figure 1**)

In the model selection, it was shown that Random Forest was the best performing machine learning classification algorithm, for both the CBC panel, and the RNA-expression panel. Furthermore, the ranked accuracy for each algorithm was also duplicated between datasets. For the RNA-expression panel, Support Vector Machines performed somewhat better with the RNA expression data than with the CBC results. Linear Discriminant Analysis was the poorest performing algorithm, in the case of RNA-expression was not able to complete (based on the high number of duplicated expression values resulting from the balancing re-sampling).

**Figure 1.**
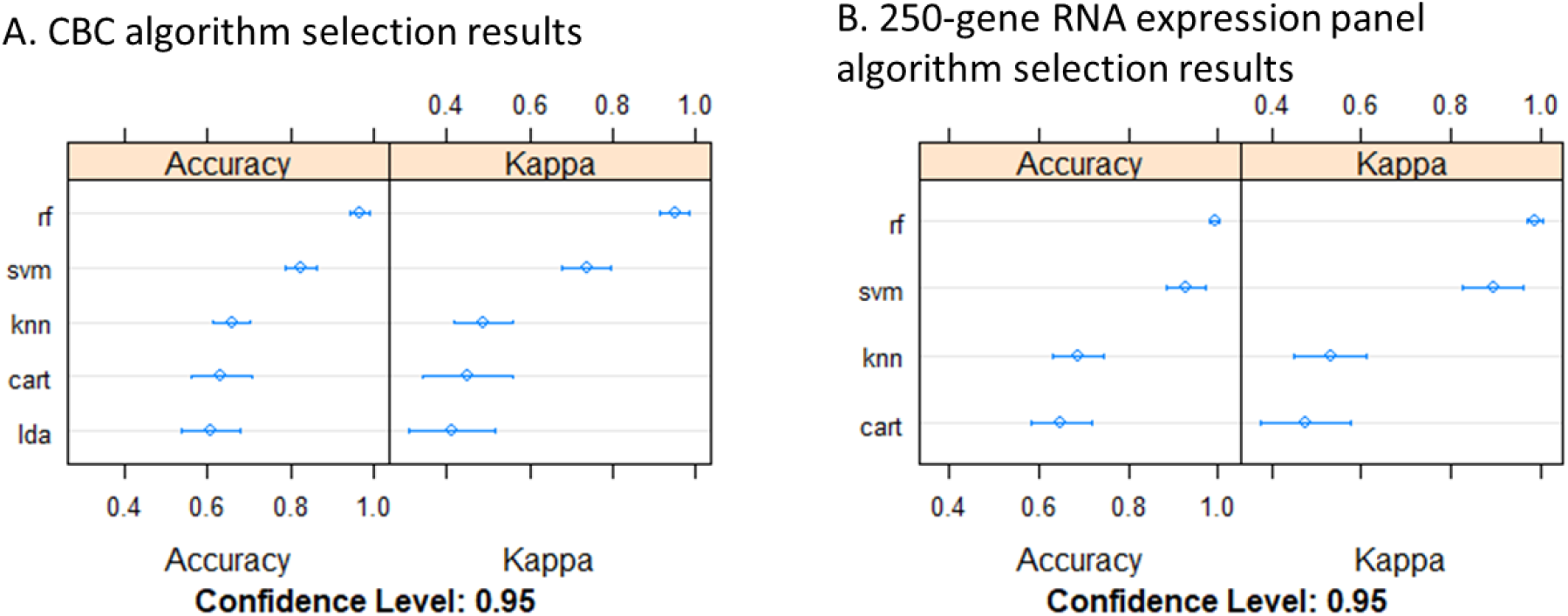
Algorithm selection results. rf = random forest, svm = support vector machines, knn = k nearest neighbors, cart = classification and regression training, lda = linear discriminant analysis.

### Performance of Random Forest **(Table 1)**

While both datasets performed reasonably well under random forest, the RNA expression panel showed nearly perfect accuracy.

**Table 1.** Confusion matrix and statistics for RNA expression VS CBC-based random forest models.

| Confusion Matrix and Statistics - RNA expression panel |  |  |  | Confusion Matrix and Statistics - Complete Blood Count |  |  |  |
| --- | --- | --- | --- | --- | --- | --- | --- |
| RNA expression | Reference |  |  | CBC | Reference |  |  |
| Prediction | Control | IBSC | IBSD | Prediction | Control | IBSC | IBSD |
| Control | 80 | 0 | 0 | Control | 18 | 0 | 0 |
| IBSC | 0 | 80 | 0 | IBSC | 1 | 20 | 0 |
| IBSD | 0 | 0 | 80 | IBSD | 1 | 0 | 20 |
| Overall Statistics | RNA expression |  | CBC |  |  |  |  |
| Accuracy | 1 |  | 0.9667 |  |  |  |  |
| 95% CI | (0.9847, 1) |  | (0.8847, 0.9959) |  |  |  |  |
| No Information Rate | 0.3333 |  | 0.3333 |  |  |  |  |
| P-Value [acc > NIR] | 2.20E-16 |  | 2.20E-16 |  |  |  |  |
| Kappa | 1 |  | 0.95 |  |  |  |  |
| Statistics by Class | RNA expression |  |  | CBC |  |  |  |
|  | Control | IBSC | IBSD | Control | IBSC | IBSD |  |
| Sensitivity | 1 | 1 | 1 | 0.9 | 1 | 1 |  |
| Specificity | 1 | 1 | 1 | 1 | 0.975 | 0.975 |  |
| Pos Pred Value | 1 | 1 | 1 | 1 | 0.9524 | 0.9524 |  |
| Neg Pred Value | 1 | 1 | 1 | 0.9524 | 1 | 1 |  |
| Prevalence | 0.3333 | 0.3333 | 0.3333 | 0.3333 | 0.3333 | 0.3333 |  |
| Detection Rate | 0.3333 | 0.3333 | 0.3333 | 0.3 | 0.3333 | 0.3333 |  |
| Detection Prevalence | 0.3333 | 0.3333 | 0.3333 | 0.3 | 0.35 | 0.35 |  |
| Balanced Accuracy | 1 | 1 | 1 | 0.95 | 0.9875 | 0.9875 |  |

In the CBC-derived random forest model, the error-rate for classifying control cases remains elevated regardless of the tree number, while in the RNA-expression model, the error converges to minimum in all categories at about 150 trees. In both cases, strict classification of Control cases was somewhat more error-prone than for IBS-D and IBS-C classifications. **(Figure 2)**

**Figure 2:**
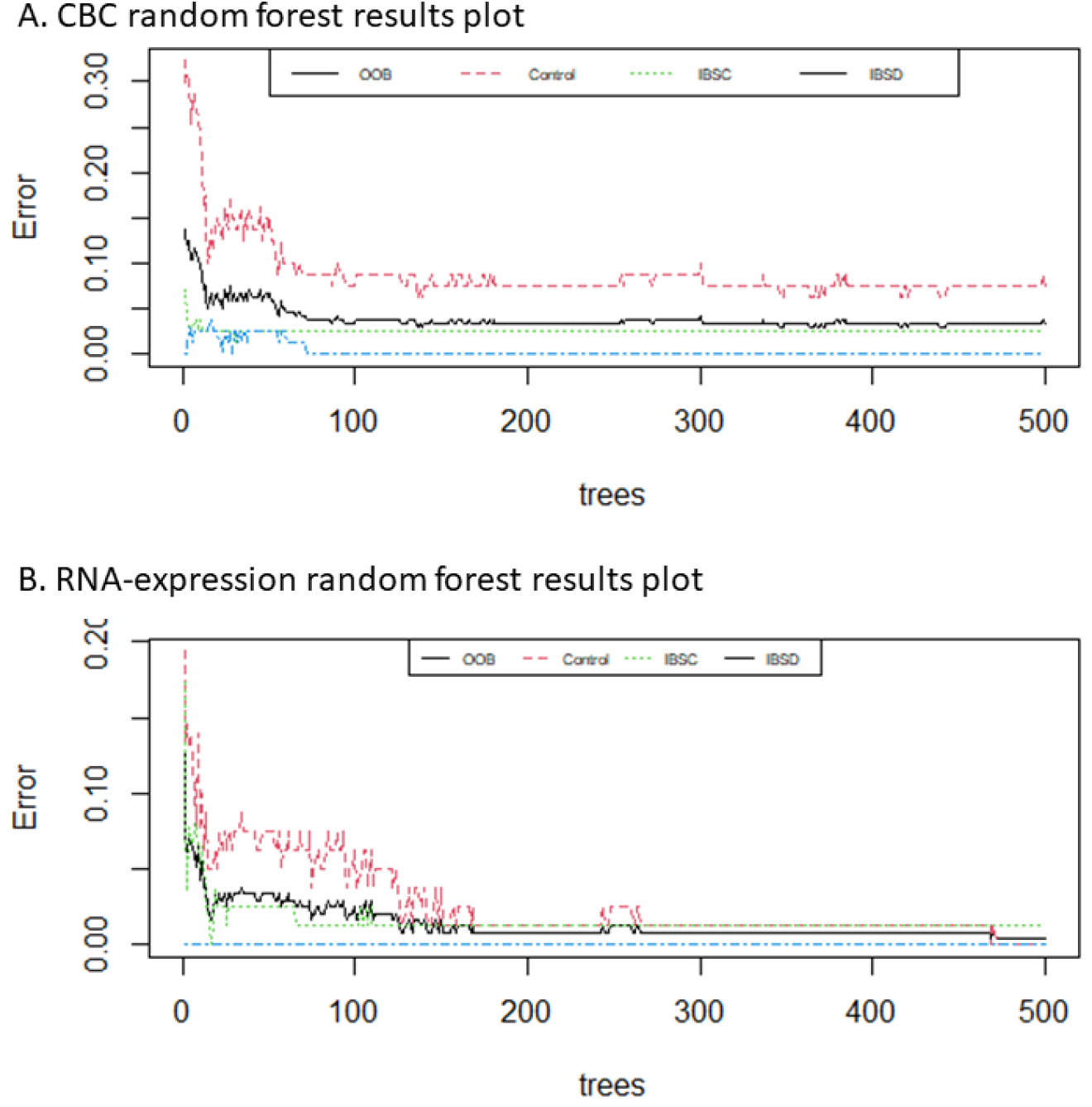
Random Forest error-rate plots.

## Discussion

A likely source of error is the incompleteness of the datasets, and artefacts resulting from the re-sampling/balancing strategy. These may produce a somewhat “easy” predictive model for the smaller original groups (IBS-D, and IBS-C), due to the duplication of rows in the dataset. Another problem faced with the interpretation of random forest models, is that the tree-selection process can be somewhat opaque. In future studies, research will further investigate trees contributing to the random forest model, and determine some of the specific, conserved decision-making nodes. This may provide some insight into the functional/biological characteristics of the models themselves.

This study shows that smaller collections of data such as those found in the CBC-D results, or small-to-medium sized RNA expression panels, can provide a highly accurate diagnostic model for traditionally symptom-based diagnoses of GI/abdominal pain. Future studies will utilize a more streamlined data intake approach to lever cloud-based data intake. Data will preferably take advantages of large cohort studies such as Framingham Heart Study (https://framinghamheartstudy.org/), or Healthy Aging in Neighborhoods of Diversity across the Life Span (https://handls.nih.gov), and utilization of Electronic Health Records (EHRs) and interoperability frameworks.

## Data Availability

This analysis utilizes previously published, open-source data from pre-print articles and the NCBI GEO database. All sources of data are linked to in the text.

https://www.ncbi.nlm.nih.gov/geo/query/acc.cgi?acc=GSE124549

https://www.ncbi.nlm.nih.gov/geo/query/acc.cgi?acc=GPL25996

https://github.com/PhyloGrok/AnalyzeBloodwork

https://www.preprints.org/manuscript/201912.0180/v1

https://www.biorxiv.org/content/10.1101/608208v1.supplementary-material

## RNA expression datasets

ImmunoGC custom Nanostring probe panel. 2019. https://www.ncbi.nlm.nih.gov/geo/query/acc.cgi?acc=GPL25996. Human buffy coat gene expression, custom 250-plex Nanostring panel. GSE124549. 2019. https://www.ncbi.nlm.nih.gov/geo/query/acc.cgi?acc=GSE124549.

## Funding

This project was partially funded from NSF XSEDE Educational Allocation to Jeffrey Robinson: “Bioinformatics Training for Applications in Translational and Molecular Biosciences”. Extreme Science and Engineering Discovery Environment (XSEDE), supported by National Science Foundation grant number ACI-1548562.

This research was developed during the course of Robinson’s faculty responsibilities at UMBC’s Translational Life Science Technology BS program, and parts of the ‘AnalyzeBloodwork’ GitHub repository have been used for training students in R-coding and methods of statistical analysis in the courses BTEC330 (Software Applications), and BTEC495 (independent student research).

## Acknowledgements

UMBC undergraduate student Mr. Daniel Gidron performed code testing as a BTEC495 intern.

## Notes

### Competing Interest Statement

The authors have declared no competing interest.

### Author Declarations

This is a re-analysis of previously published data, the full links and references to the previously published, open-sourced data have been included in the text.

## Citations

1. Manning AP., et al. Towards positive diagnosis of the irritable bowel. Br Med J. 2(6138):653–4.

2. Van Leeuwen AM, Kranpitz TR, Smith L. 2006. Complete Blood Count. pp. 413–420 In: Davis’s Comprehensive Handbook of Laboratory and Diagnostic Tests with Nursing Implications 2^nd^. F.A. Davis Company, Philadelphia.

2. Robinson JM., et al. 2019. Complete blood count with differential: An effective diagnostic for IBS subtype in the context of BMI? BioRxiv. doi: https://doi.org/10.1101/608208.

3. Robinson J. 2019. Differential Gene Expression Associated with BMI, Gender, and IBS-subtype in Human White Blood Cells: Results from a Custom 250-plex Nanostring Probe Panel. Preprints 2019, 2019120180 (doi: 10.20944/preprints201912.0180.v1).

4. Brownlee. 2016. Your First Machine Learning Project in R Step-By-Step. https://machinelearningmastery.com/machine-learning-in-r-step-by-step/.

5. Brownlee. 2020. Machine Learning Mastery With R: Discover The Most Popular Machine Learning Platform With Step-By-Step Tutorials And End-To-End Projects v1.1. Copyright Jason Brownlee, Machine Learning Mastery with R.

6. Kuhn M. 2021. caret: Classification and Regression Training. R package version 6.0-88. https://CRAN.R-project.org/package=caret.

7. Yiu, T. 2019. Understanding Random Forest: How the algorithm works and why it is so effective. https://towardsdatascience.com/understanding-random-forest-58381e0602d2.

8. Robinson, J. 2021. AnalyzeBloodwork1.5. https://github.com/PhyloGrok/AnalyzeBloodwork. doi: https://doi.org/10.5281/zenodo.5348283.

