## Supplementary Files 1 for "Predictive Classification of IBS-subtype: Performance of a 250-gene RNA expression panel vs. Complete Blood Count (CBC) profiles under a Random Forest model": IBSclassification.R

```
## IBSclassification.R
## Machine Learning for predictive classification of IBS subtype
## Comparison between Complete Blood Count (CBC) and a 250-gene RNA expression panel

## A. SETUP ENVIRONMENT

## A1. SET WORKING DIRECTORY
if (!require("rstudioapi")) {
  install.packages("rstudioapi", dependencies = TRUE)
  library(rstudioapi)
}

setwd(dirname(getActiveDocumentContext()$path))

## A2. Check for installed packages
packagelist <- c("mime", "groupdata2","shiny", "rstudioapi", "dplyr", "tidyr", "knitr", "caret", "e1071", "ellipse", "ggplot2", "scatterplot3d", "ggvis")
not_installed <- packagelist[!(packagelist %in% installed.packages()[ , "Package"])]    # Extract not installed packages
if(length(not_installed)) install.packages(not_installed)       

## Load packages
invisible(lapply(packagelist, library, character.only = TRUE))

## B. Load CBC data
IBS1 <- read.csv("../data/RobinsonEtAl_Sup1.csv", header = TRUE)
IBS1$ID <- as.character(IBS1$ID)

## Generate subset list
IBSlist <- c("ID", "IBS.subtype")
CBClist <- c("ID", "IBS.subtype","BMI", "Monocytes", "Lymphocytes", "Neutrophils", "Basophils", "Eosinophils", "RBC", "ESR", "MCH", "HCT", "PlateletCount", "MPV")

## Subset data by list
IBS <- IBS1[,IBSlist]
CBC <- IBS1[,CBClist]

## C. CBC - IMPUTE MISSING VALUES USING CALCULATED SAMPLE MEANS

## Generate a list of columns containing NA values
list_na <- colnames(CBC)[ apply(CBC, 2, anyNA) ]
list_na

for (j in list_na){
  print(j)
}

## CALCULATE MEANS
average_missing <- apply(as.matrix(CBC[,colnames(CBC) %in% list_na]), 
                         2, 
                         mean, 
                         na.rm=TRUE)
average_missing

## REPLACE NA VALUES with the calculated means, for each subset
CBC.replacement <- CBC

sum(is.na(CBC.replacement$i))

CBC.replacement <- CBC.replacement %>%
  mutate(
    BMI = ifelse(is.na(BMI), average_missing[1], BMI),
    Monocytes = ifelse(is.na(Monocytes), average_missing[2], Monocytes),
    Lymphocytes = ifelse(is.na(Lymphocytes), average_missing[3], Lymphocytes),
    Neutrophils = ifelse(is.na(Neutrophils), average_missing[4], Neutrophils),
    Basophils = ifelse(is.na(Basophils), average_missing[5], Basophils),
    Eosinophils = ifelse(is.na(Eosinophils), average_missing[6], Eosinophils),
    RBC = ifelse(is.na(RBC), average_missing[7], RBC),
    ESR = ifelse(is.na(ESR), average_missing[8], ESR),
    MCH = ifelse(is.na(MCH), average_missing[9], MCH),
    HCT = ifelse(is.na(HCT), average_missing[10], HCT),
    PlateletCount = ifelse(is.na(PlateletCount), average_missing[11], PlateletCount),
    MPV = ifelse(is.na(MPV), average_missing[12], MPV)
      )

## D. BALANCING - Balance IBS groups to have 100/group by re-sampling
## Probably better to do this by using synthetic method based on distribution

## Unbalanced group sizes
CBC.replacement %>%
  count(IBS.subtype) %>%
  kable(align = 'c')

## Balance by making each group have 100 rows
CBC_balanced <- balance(CBC.replacement, 100, cat_col = "IBS.subtype") %>% 
  arrange(IBS.subtype)

## Post-balancing
CBC_balanced %>%
  count(IBS.subtype) %>%
  kable(align = 'c')

# Summarize group proportions
percentage <- prop.table(table(CBC_balanced$IBS.subtype)) * 100
cbind(freq=table(CBC_balanced$IBS.subtype), percentage=percentage)

# Summarize attribute distributions
summary(CBC_balanced)

## E. PREDICTIVE CLASSIFICATION MODELS FOR IBS-SUBTYPES 
## Derived from https://machinelearningmastery.com/machine-learning-in-r-step-by-step/

CBCdf <- CBC_balanced[ -c(1) ]

## E1. IDENFITY THE BEST-PERFORMING PREDICTIVE CLASSIFICATION ALGORITHM 

# create a list of 80% of the rows in the original dataset we can use for training
validation_index <- createDataPartition(CBCdf$IBS.subtype, p=0.80, list=FALSE)
# select 20% of the data for validation
validation <- CBCdf[-validation_index,]
# use the remaining 80% of data to training and testing the models
CBCdf <- CBCdf[validation_index,]

## E1a. Set validation parameters
control <- trainControl(method="cv", number=10)
metric <- "Accuracy"

## E1b. Generate models from a selection of classification algorithms.
# a) linear algorithms
set.seed(7)
fit.lda <- train(IBS.subtype~., data=CBCdf, method="lda", metric=metric, trControl=control)
# b) nonlinear algorithms
# CART
set.seed(7)
fit.cart <- train(IBS.subtype~., data=CBCdf, method="rpart", metric=metric, trControl=control)
# kNN
set.seed(7)
fit.knn <- train(IBS.subtype~., data=CBCdf, method="knn", metric=metric, trControl=control)
# c) advanced algorithms
# SVM
set.seed(7)
fit.svm <- train(IBS.subtype~., data=CBCdf, method="svmRadial", metric=metric, trControl=control)
# Random Forest
set.seed(7)
fit.rf <- train(IBS.subtype~., data=CBCdf, method="rf", metric=metric, trControl=control)

## E2. IDENTIFY THE BEST-PERFORMING PREDICTIVE CLASSIFICATION ALGORITHM
CBCresults <- resamples(list(lda=fit.lda, cart=fit.cart, knn=fit.knn, svm=fit.svm, rf=fit.rf))
summary(CBCresults)

dotplot(CBCresults)

# Export dotplot with model validation results
png("../fig_output/CBCclassification.png")
H1 <- dotplot(CBCresults)
print(H1)
dev.off()

## E3. Estimate 'skill' of models with confusion matrix
predictions <- predict(fit.rf, validation)
confusionMatrix(predictions, as.factor(validation$IBS.subtype))

plot(fit.rf)
plot(fit.rf$finalModel)


## Provide a legend for the error rate plot
## https://stats.stackexchange.com/questions/51629/multiple-curves-when-plotting-a-random-forest
fit.rf.legend <- {colnames(fit.rf$finalModel$err.rate)}

legend("top", cex=0.5, legend=fit.rf.legend, lty=c(1,2,3), col=c(1,2,3), horiz=T)


## F. RNA expression for classification of IBS subtype
## Load RNA expression data (Nanostring normalized counts)

## F1a. Import Nanostring Normalized Count data
mRNAexp <- read.csv("../data/mRNAnorm.csv", header = TRUE)
mRNAexp$ID <- as.character(mRNAexp$ID)

## F1b. Merge datasets and remove NA rows
##  Note - Values will not be imputed for IDs with no data
IBS_RNA <- full_join(IBS, mRNAexp, by = "ID")
IBS_RNA <- na.omit(IBS_RNA)

## F2. BALANCE RNA dataset - Balance IBS groups to have 100/group by re-sampling

## Unbalanced group sizes
IBS_RNA %>%
  count(IBS.subtype) %>%
  kable(align = 'c')

## Balance by making each group have 100 rows
IBS_RNA <- balance(IBS_RNA, 100, cat_col = "IBS.subtype") %>% 
  arrange(IBS.subtype)

## Post-balancing
IBS_RNA %>%
  count(IBS.subtype) %>%
  kable(align = 'c')

## F3. IDENFITY THE BEST-PERFORMING PREDICTIVE CLASSIFICATION ALGORITHM 
RNAdf <- IBS_RNA[ -c(1) ]

## F3a. Split validation and training sets
# create a list of 80% of the rows in the original dataset we can use for training
validation_index <- createDataPartition(RNAdf$IBS.subtype, p=0.80, list=FALSE)
# select 20% of the data for validation
validation <- RNAdf[-validation_index,]
# use the remaining 80% of data to training and testing the models
RNAdf <- RNAdf[validation_index,]

## F3b. Set validation parameters
control <- trainControl(method="cv", number=10)
metric <- "Accuracy"

## F3c. Generate models from a selection of classification algorithms.
# a) linear algorithms  ## Note LDA 
set.seed(7)
fit.lda <- train(IBS.subtype~., data=RNAdf, method="lda", metric=metric, trControl=control)

# b) nonlinear algorithms
# CART
set.seed(7)
fit.cart <- train(IBS.subtype~., data=RNAdf, method="rpart", metric=metric, trControl=control)
# kNN
set.seed(7)
fit.knn <- train(IBS.subtype~., data=RNAdf, method="knn", metric=metric, trControl=control)
# c) advanced algorithms
# SVM
set.seed(7)
fit.svm <- train(IBS.subtype~., data=RNAdf, method="svmRadial", metric=metric, trControl=control)
# Random Forest
set.seed(7)
fit.rf <- train(IBS.subtype~., data=RNAdf, method="rf", metric=metric, trControl=control)

## F4. IDENTIFY THE BEST-PERFORMING PREDICTIVE CLASSIFICATION ALGORITHM
RNAresults <- resamples(list(cart=fit.cart, knn=fit.knn, svm=fit.svm, rf=fit.rf))
summary(RNAresults)

dotplot(RNAresults)

# Export dotplot with model validation results
png("../fig_output/RNAclassification.png")
H1 <- dotplot(RNAresults)
print(H1)
dev.off()

# F5. Estimate 'skill' of Random Forest with confusion matrix
predictions <- predict(fit.rf, validation)
confusionMatrix(predictions, as.factor(validation$IBS.subtype))

## G. Plot the results for a CBC vs RNA comparison
## https://stackoverflow.com/questions/39636186/plot-decision-tree-in-r-caret

plot(fit.rf)
plot(fit.rf$finalModel)

## Provide a legend for the error rate plot
## https://stats.stackexchange.com/questions/51629/multiple-curves-when-plotting-a-random-forest
fit.rf.legend <- {colnames(fit.rf$finalModel$err.rate)}

legend("top", cex=0.5, legend=fit.rf.legend, lty=c(1,2,3), col=c(1,2,3), horiz=T)

## Visualize the shortest-length trees for each of the parameters
## https://shiring.github.io/machine_learning/2017/03/16/rf_plot_ggraph
```
