## Supplementary figures and images for "Predictive Classification of IBS-subtype: Performance of a 250-gene RNA expression panel vs. Complete Blood Count (CBC) profiles under a Random Forest model"

### CBCclassification.png

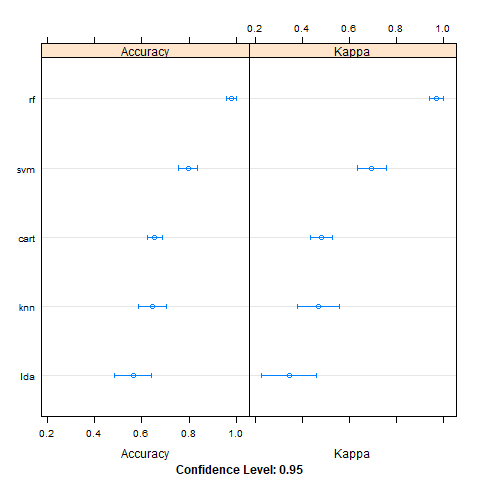

### RNAclassification.png

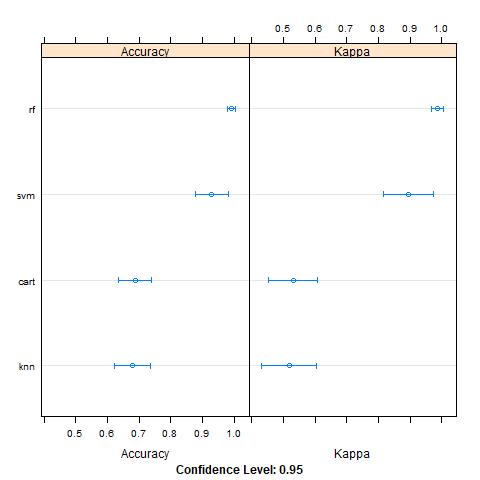
